# Immune, metabolic, anatomical, and functional features of people after successful tuberculosis treatment: an exploratory analysis

**DOI:** 10.1101/2025.01.30.25321373

**Authors:** Tariq Webber, Candice Macdonald, Michele Tameris, Nicolette Tredoux, Anandi Bierman, Andrea Gutschmidt, Susanne Tönsing, Andriëtte Hiemstra, Firdows Noor, Candice Snyders, Tracy Richardson, Bernadine Fransman, Brian Allwood, Mark Hatherill, Leanie Kleynhans, André G. Loxton, Gerhard Walzl, Novel Chegou, Nelita du Plessis, Jane A Shaw, Stephanus T. Malherbe

## Abstract

**Background:** We explored the underlying mechanisms that may drive Post-tuberculosis (TB) lung disease, a multifactorial, heterogenous, and prevalent disease.

**Methods:** Extensive clinical phenotyping through fluorine-18 fluorodeoxyglucose (FDG) positron emission tomography (PET)-computed tomography (CT) scans, pulmonary function testing, and symptom and quality of life questionnaires, was performed on a cohort of 48 adults who completed TB treatment within 6 months prior. Immunological characteristics of paired blood- and bronchoalveolar lavage fluid (BALF)-derived immune cells were assessed by multiplex bead-based immunoassay, ELISA and flow cytometry.

**Results:** There was agreement between measures of inflammation on PET, the severity of anatomical abnormalities on CT, and pulmonary function testing. However, of these, only PET was associated with exercise tolerance and symptom scores. Measures of radiologic extent (total glycolytic activity and SUVmax on PET, and segments involved on CT) also correlated with proteins detected in blood that implicate Type 1 (IFN-γ, TNFα, IL-12) and Type 2 (IL-4, IL-33) responses, ongoing remodelling of lung tissue (MMPs), airways and vasculature (VEGF), as well as subsets of activated CD8^+^ and CD4^+^ T-cells.

**Conclusion:** The radiologic extent of structural post-TB lung involvement is associated with a range of impaired lung function measures and immunological dysregulation. Our findings suggest that obstructive and restrictive lung pathology due to pulmonary tuberculosis do not occur in opposition but rather point towards a mixed pathology in most TB survivors.

## INTRODUCTION

The estimated 155 million survivors of tuberculosis (TB) are affected by potentially lifelong consequences of post-TB lung disease (PTLD).^1^ Up to 75% have a pulmonary function test abnormality after treatment, in any of restrictive, obstructive or mixed patterns.^2^ The most common trajectory is a slight improvement of function for up to 12 months after treatment, at which point the dysfunction becomes stagnant. However, in some cases further deterioration occurs, especially in parameters measuring gas trapping.^3^ Most survivors also have residual lung lesions on imaging with ongoing inflammation detectable on ^18F^fluorodeoxyglucose positron-emission tomography-computed tomography (FDG PET-CT).^4, 5^ Post-TB lung disease (PTLD) presents with a broad clinical spectrum of severity.^2,6,7^ People post-TB may be asymptomatic, or experience severe disability that contributes to the TB-associated disability burden.^8^

While there is a growing body of evidence and awareness of PTLD, little is known about the underlying mechanisms driving this multifactorial and heterogenous condition, with no established standard-of-care interventions despite a high burden of disease.^9^

The aim of the study was to better understand the spectrum of pathophysiological processes associated with the different clinical features of PTLD. We therefore performed extensive clinical phenotyping of adults early after successful TB treatment completion using FDG PET-CT, full pulmonary function testing, and validated symptom and quality of life questionnaires. We performed bronchoscopy with bronchoalveolar lavage (BAL) and used a multiplex bead-based immunoassay and flow cytometry on both BAL fluid (BALF) and peripheral blood to compare the patterns of key proinflammatory and regulatory cytokines, stress hormones, and immune cell subtypes between patients according to the key clinical disease markers on imaging and pulmonary function tests.

## METHODS

### Recruitment and clinical procedures

We recruited sequential consenting adults who were within 6 months of the end of successful treatment for drug sensitive TB, at primary healthcare facilities in the Western Cape, South Africa. TB cure was confirmed by negative sputum liquid culture (BACTEC MGIT, Becton Dickinson, USA) and clinical assessment at enrolment. We excluded those who recently used systemic corticosteroids, had current active infection other than virally-suppressed HIV, with known or suspected malignancy, were pregnant or breast-feeding. All participants gave written informed consent. The study was approved by the Stellenbosch University Health Research Ethics Committee and the University of Cape Town Human Research Ethics Committee.

FDG PET-CT scans were read by a nuclear medicine physician, specialist radiologist and independently by an investigator with PET-CT expertise. CT-based measurements in the lungs included 1) cavity volumes, 2) volumes of very high-density lung lesions corresponding with TB lesions and fibrosis using a −100 Hounsfield Unit (HU) threshold,^10^ 3) very low-density volumes that include emphysema and gas trapping using a threshold of >-910 Hounsfield Units (HU).^11^ We grouped lung bronchopulmonary segments that are commonly co-affected, namely: S1&2, S3, S4&S5, S6, and S7-10 for left and right respectively and captured the presence of morphological lesion types per segment group as qualitative assessment. PET-based lung measurements were total glycolytic activity (TGA) and most intense uptake in the lung (SUVmax).

Bronchoscopy with 240 mL bronchoalveolar lavage was performed under conscious sedation.^6^ We directed the bronchoscope to airways with proximity to metabolically active lesions where possible.

All participants completed the Chronic Obstructive Pulmonary Disease (COPD) Assessment Test (CAT) and the St George’s Respiratory Questionnaire (SGRQ).^12–14^ A six-minute walking test (6MWT) was conducted on a 30 m flat indoor surface,^15^ and the five-times-sit-to-stand test (5TSTS) by recording the time in seconds that the participant needed to stand up and sit down five times.^16^ Participants had spirometry with routine measurements including forced expiratory volume in 1 second (FEV1) and forced vital capacity (FVC), diffusing capacity of the lungs for carbon monoxide (D_L_CO), and body plethysmography in an accredited lung function laboratory according to international guidelines and standards. GLI reference equations were used.

### Laboratory procedures

We performed Luminex® and ELISA assays on serum and BAL fluid. We designed a targeted panel of analytes of interest in lung disease processes (fibrosis, cavitation, emphysema, bronchiectasis etc), and proinflammatory and regulatory immune responses (Supplementary Table 1). BALF specimens were filter-sterilized, freeze dried, and resuspended in PBS to concentrate the specimens to 5:1.

Peripheral blood mononuclear cells (PBMCs) were isolated from venous blood collected in heparinized tubes. A flow cytometric panel was designed for the Cytek Aurora flow cytometer, including 21 lymphocyte cell surface proteins and intracellular cytokines (Supplementary Table 2), for the identification of subsets and activation states of CD4 Helper T cells, Cytotoxic T cells, and B cells. For detailed methods see Supplementary Materials.

### Statistical analysis

Analysis was conducted with R version 4.3.3 [R Core Team 2023]. Data were tested for normality using Shapiro-Wilks test. All variables were analysed using the non-parametric Kendall’s Tau method during correlation analyses due to the majority having non-normal distributions. We report the significant correlations (p < 0,05) after correction for confounders such as age, BMI, years smoking and illicit drug use (Supplementary Table 3). The values of serum analytes in BAL were normalised to the Albumin levels in the same sample to correct for the dilution factor. To minimise the impact of multiple testing, significant correlations were interrogated using sensitivity analysis, indicating instances in which one or more subsets of 5% of the data drive a previously significant correlation, or the effect does not hold true in a reduced sample size.

## RESULTS

We screened 67 adults within 6 months of TB treatment, of whom 48 met inclusion criteria and underwent FDG PET-CT scans, spirometry, respiratory questionnaires, and exercise tolerance tests (6MWT and 5TSTS), at a median 62 days after TB treatment completion (range 4–160 days). Due to challenges with equipment (gas stock-out) or participant compliance, acceptable plethysmography and D_L_CO could only be acquired in 31 participants. 40 participants underwent BAL, seven were excluded due to safety concerns, and one lost to follow up. Supplementary figure 1 shows the flow of recruitment and study procedures.

### Abnormalities on pulmonary function tests and FDG PET-CT were common and highly variable after successful TB treatment

Table 1 shows demographics and other characteristics for all participants, subdivided into spirometry classes. We classified 26 (54.2%) of the participants with normal spirometry, 6 (12.5%) with an obstructive pattern, 9 (18.7%) with possible restriction, and 7 (14.6%) with a mixed pattern. Only one participant displayed significant bronchodilator response (>10% increase in FEV1 or FVC). Impaired forced mid-expiratory flow (FEF25-75) was found in 23 (48%) of the participants, and distributed in all spirometry class groups, including 15% of those with FVC and FEV1 within the normal range, 83% in the obstructive group, 78% in the restrictive group and all in the mixed group (Supplementary Table 4a). Seven of the 23 (30.4%) showed a >10% improvement in FEF25-75 with bronchodilation. 17 of 31 (55%) had some diffusion abnormality mostly attributable to pulmonary vascular abnormality (Supplementary Table 4b). Abnormalities on plethysmography or D_L_CO were detected in ten of the 26 (38.5%) participants with spirometry in the normal range.

**Table 1:**
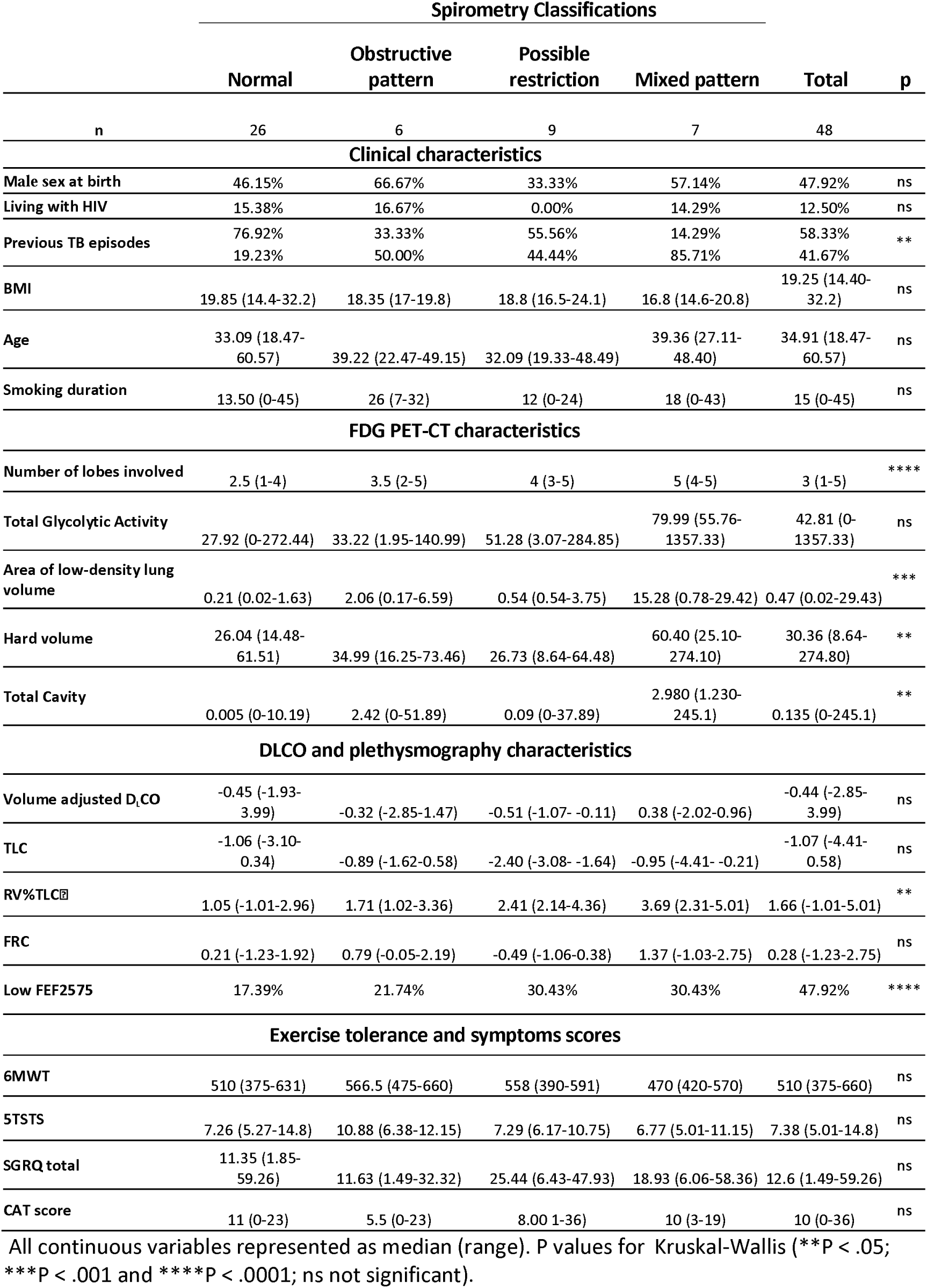
Demographic, FDG PET-CT scan, and pulmonary function test characteristics for all participants by spirometry pattern post-TB.

On CT, all participants had lesion(s) attributable to the aftermath of TB. Residual FDG avidity on PET was present in 41 (85.4%), defined by total glycolytic activity (TGA) >5 or SUVmax higher than mild (compared to aortic blood pool as reference). There was a wide range of morphological features, usually co-occurring in an individual (Figure 1). Small nodules were the most common, affecting 31% of segment groups, followed by fibrotic scarring (27%), cavitation (16%), large nodules (15%), bronchiectasis (12%), bullae (10%), and consolidation (4%). Differences in the gross pathology between the groups were not as clear as expected, with adjacent lobes and segments often reflecting contrasting fates; and areas of low density often surrounding dense inflammatory or fibrotic lesions.

**Figure 1:**
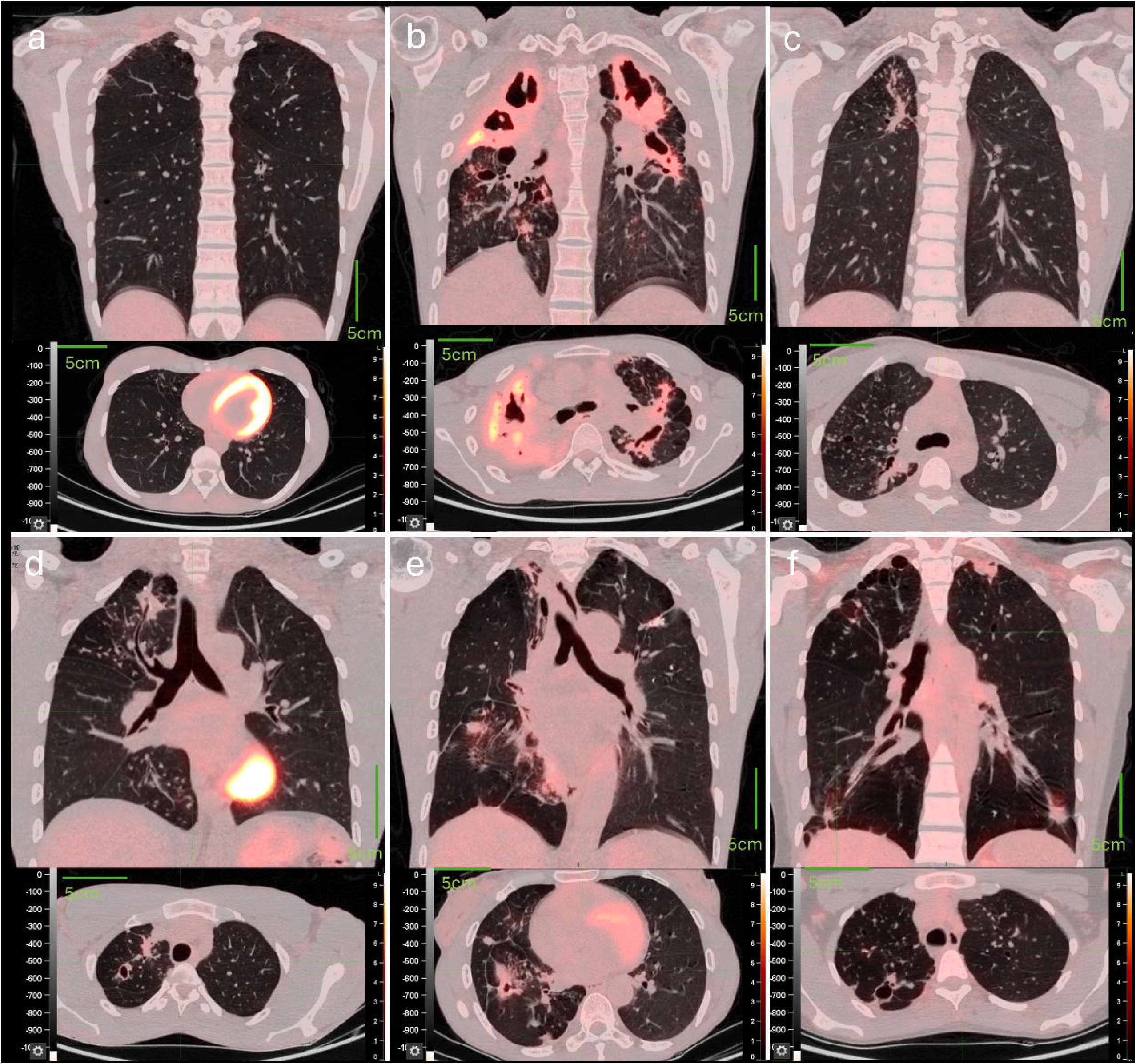
Transverse and sagittal views of CT images with PET overlay. Fig 1a: A participant with no FDG-avid lesions in lungs, but some small nodules and linear fibrotic lesions in the right apex and right middle lobe. Fig 1b: A participant with the most residual glycolytic activity on PET. The image shows large areas of destruction, cavitation, fibrosis, and nodules – especially in the upper lobes. Fig 1c: A participant with median FEV1 value in the normal spirometry (FEV1 and FVC normal) category. The right upper lobe shows diffuse small nodules and posterior dense lesions with calcifications, as well as decreased density of lung tissue. Fig 1d: A participant with the median FEV1 value in the obstructive category (low FEV1, normal FVC) on spirometry. The right upper lobe shows numerous small nodules, cavitation, and an apical dense lesion with calcifications, as well as decreased density lung tissue; the right lower lobe shows numerous small nodules. Fig 1e: A participant with the median FEV1 value in the restrictive category (low FEV1 and low FVC and normal FEV1/FVC ratio) on spirometry. There are dense lesions and loss of volume in right upper and lower lobes, with heterogenous pathology in other lobes. Fig 1f: A participant with the median FEV1 value in the mixed category (low FEV1, low FVC with low FEV1/FVC ratio) on spirometry. There are emphysematous changes and cavitation in right upper lobe, with heterogenous pathology in other lobes.

### There was overall agreement on severity of disease between FDG PET-CT and pulmonary function testing

The mixed spirometry class had the worst median for several severity of disease indicators as shown in Table 1, including: 1) the lowest BMI, 2) the highest number of lobes with any lesions in keeping with previous TB on CT, 3) the largest cavity volume, 4) the largest volume of high-density lesions (>-100 HU), as well as 5) total glycolytic activity on PET. The obstructive group had the longest median smoking duration and lungs with the highest volume of low-density lungs on CT (<-910HU).

Quantified image metrics correlated with pulmonary function test results (Figure 2a) as follows: 1) Volume of low density to FEV1, FEF2575, and Residual Volume/Total lung capacity ratio – all indicative of large or small airway obstruction or gas trapping, 2) Features of inflammation, destruction, and fibrosis (respectively TGA, cavity volume, high density lesion volume) to low FVC, indicative of loss of parenchyma. TGA, cavity volume and dense lesion volume correlated more strongly with FEV1 and FEF2575; and FVC and FEV1 values were also strongly correlated (p < 0.0001). When further exploring the correlation between the lobes affected and pulmonary function as shown in Table 2, we found a trend for both the decrease in total lung capacity, as well as the increase in residual volume with every additional lobe affected.

**Figure 2a:**
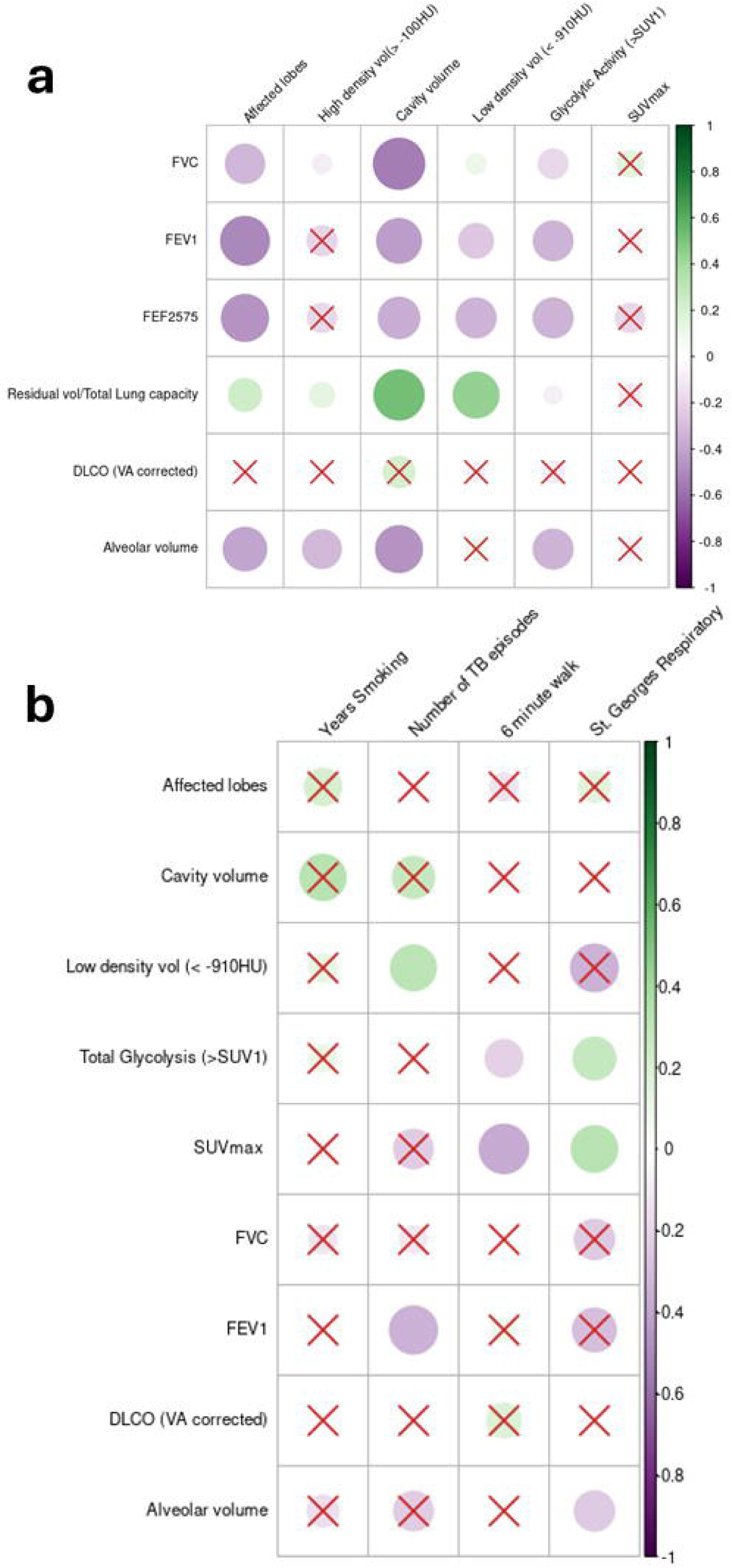
Correlation of quantified imaging variables to pulmonary function. Cross marks indicate non-significance (p ≥ 0.05). Note that increasing lung function values indicate improving function, except for residual volume/total lung capacity ratio, while high imaging values indicate more pathology, hence the inverse correlations. Figure 2b: Correlation of quantified imaging variables and lung function to effort tolerance tests, symptom scores and clinical variables.

**Table 2:**
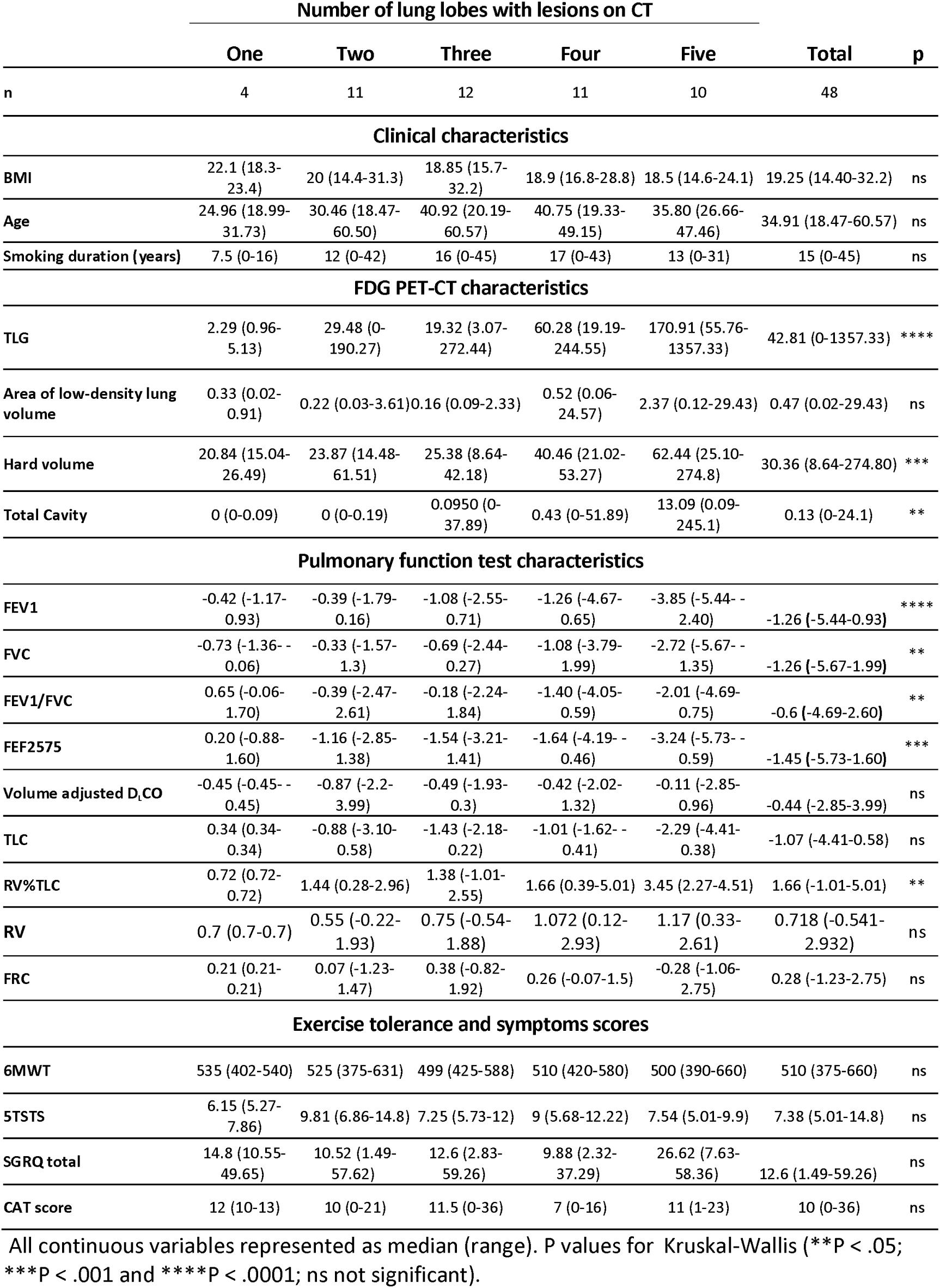
Demographic, FDG PET-CT scan, and pulmonary function characteristics for all participants, by number of lung lobes with any residual lesions post-TB.

### Exercise tolerance and quality of life after TB treatment were dissociated from imaging severity and pulmonary function testing

Most participants performed well in the exercise tests and did not indicate severe subjective disability on the questionnaires (Table 1). We found no difference in exercise tolerance (6MWT and 5TSTS), or quality-of-life questionnaires (the SGRQ and CAT COPD) between the spirometry groups. Furthermore, these measures did not correlate with conventional pulmonary function test measures of severity (FEV1, FVC) and CT characteristics, but the St Georges Respiratory Questionnaire and 6MWT was positively correlated with measures of inflammation (TGA & SUVmax) on PET (Figure 2b).

### Concentrations of proinflammatory cytokines in serum were correlated with measures of inflammation and the number of lung segments with lesions on FDG PET-CT

Correlations were observed between inflammation markers on PET (TGA or SUVmax), and both Type 1 (interferon (IFN)γ, tumour necrosis factor (TNF)α, interleukin (IL)-2, IL-12) and Type 2 (IL-4, IL-33) pro-inflammatory responses, regulatory response (IL-10), chemo-attractants (CXCL-9, CXCL-12), selected growth factors, matrix metalloproteins (MMPs) (1,8), and tissue inhibitors of MMPs (TIMPs) (1, 3) (Figure 3a). The number of affected bronchopulmonary segment groups on CT had similar association patterns, but with significant correlations to MMPs (1,8) and chemo-attractants (IL-8, CCL-2), and fewer correlations with Type 1 and Type 2 response proteins. Pulmonary function tests were not correlated with inflammatory cytokines, but worse function did correlate with levels of selected growth factors (PDGF, FGFβ), TIMP-1 and MMPs (1,8). Surfactant protein D (SP-D), which is crucial for surfactant regulation and innate immunity and may induce immune pathology,^17^ correlated with markers of severity, extent of lung involvement and worse pulmonary function. Conversely, the immunomodulatory cytokine, uteroglobin, correlated to better pulmonary function.

**Figure 3a:**
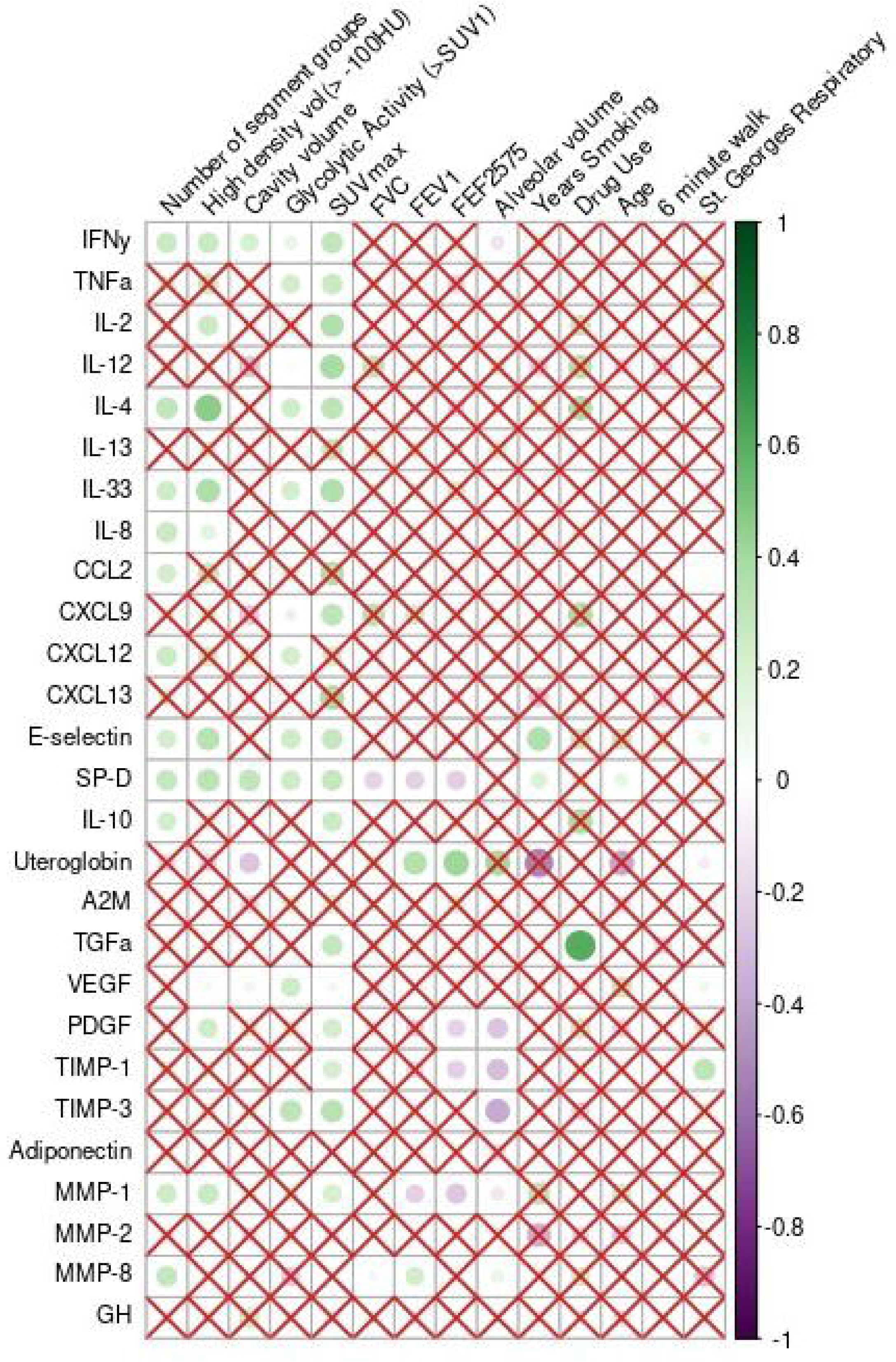
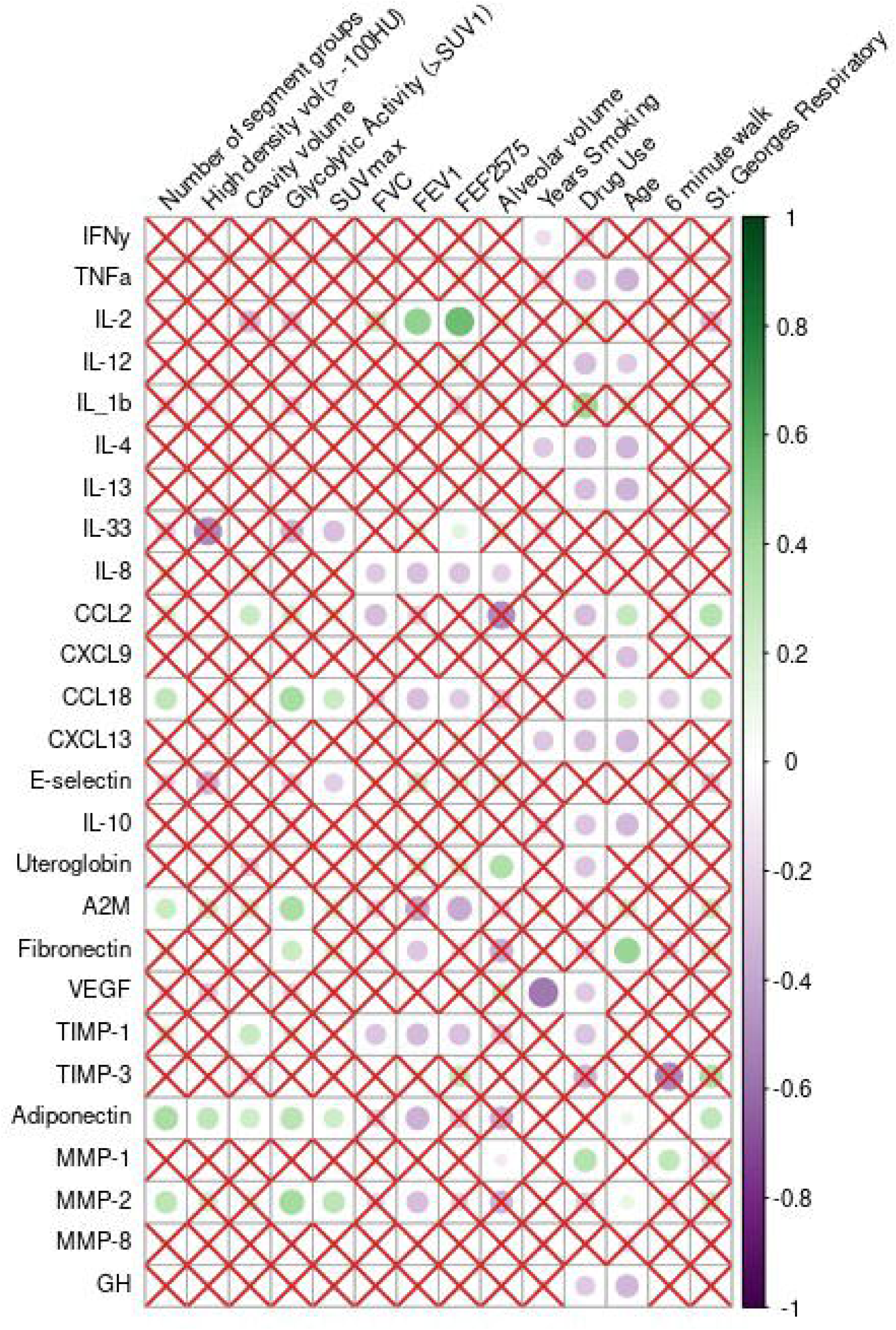
Correlation matrix of quantified imaging variables, pulmonary function tests and participant characteristics to protein factors in serum. Cross marks indicate non-significance (p ≥ 0.05). Note that increasing lung function values indicate improving function (except for residual volume/total lung capacity ratio), while high imaging values indicate more pathology, hence the inverse correlations. Figure 3b: Correlation matrix of quantified imaging variables, pulmonary function tests and participant characteristics to protein factors in bronchoalveolar lavage.

### Worsening severity of cavity volume, FDG PET inflammation, FEV1, and symptoms were correlated with higher levels of markers of leukocyte trafficking, collagen degradation and remodelling, regulation and repair in BAL

The analysis of BALF proteins was markedly influenced by age, smoking, and illicit drug use (which appeared to have a global suppressive effect) more clearly than serum proteins (Figure 3b, Supplementary Table 3), although some notable correlations remained after correction. More extensive lesions and worse lung function were correlated to lower IL-2 (in contrast to serum values), higher IL-8 and other chemo-attractants like CCL-18, levels of adiponectin, fibronectin, MMP-2, and TIMP-1. The cavity volume correlated positively with levels of BAL CCL2, TIMP-1 and adiponectin, while FEV1 was inversely correlated with IL-8, CCL18, fibronectin, TIMP-1, adiponectin, and MMP-2, and positively with IL-2 levels. The amount of glycolytic activity on FDG-PET correlated positively with CCL18, A2M, fibronectin, adiponectin, and MMP-2. Interestingly the SGRQ was positively correlated with CCL2, CCL18, and adiponectin.

### Higher measures of inflammation and the severity of lesions on FDG PET-CT correlated with reduced regulation of inflammation by T helper cells, and priming of cytotoxic T cells for migration to the site of disease

Participants with higher CT scan measures of involvement (low- and high-density volumes, cavity volume, number of segment groups involved) and higher FDG-PET avidity (TGA, and SUVmax) correlated positively with clusters of helper T cells displaying memory (CD45RA-) and effector phenotypes (CD197-), and negatively with naive T cells (CD45RA+CD197+). Additionally, these features of radiologic involvement were positively correlated(Kendall’s tau < 0.2) with percentages of CD194+ Th2 and CD183+ Th1 cells, and negatively with the presence of CD196+ Th17 subpopulations (Figure 4a). The Treg population (CD25+ CD127lo) showed a weak negative correlation with TGA. We hypothesize that this may represent reduced Treg regulation of inflammation, with downstream dysregulation of TGFβ contributing to fibrosis.

**Figure 4:**
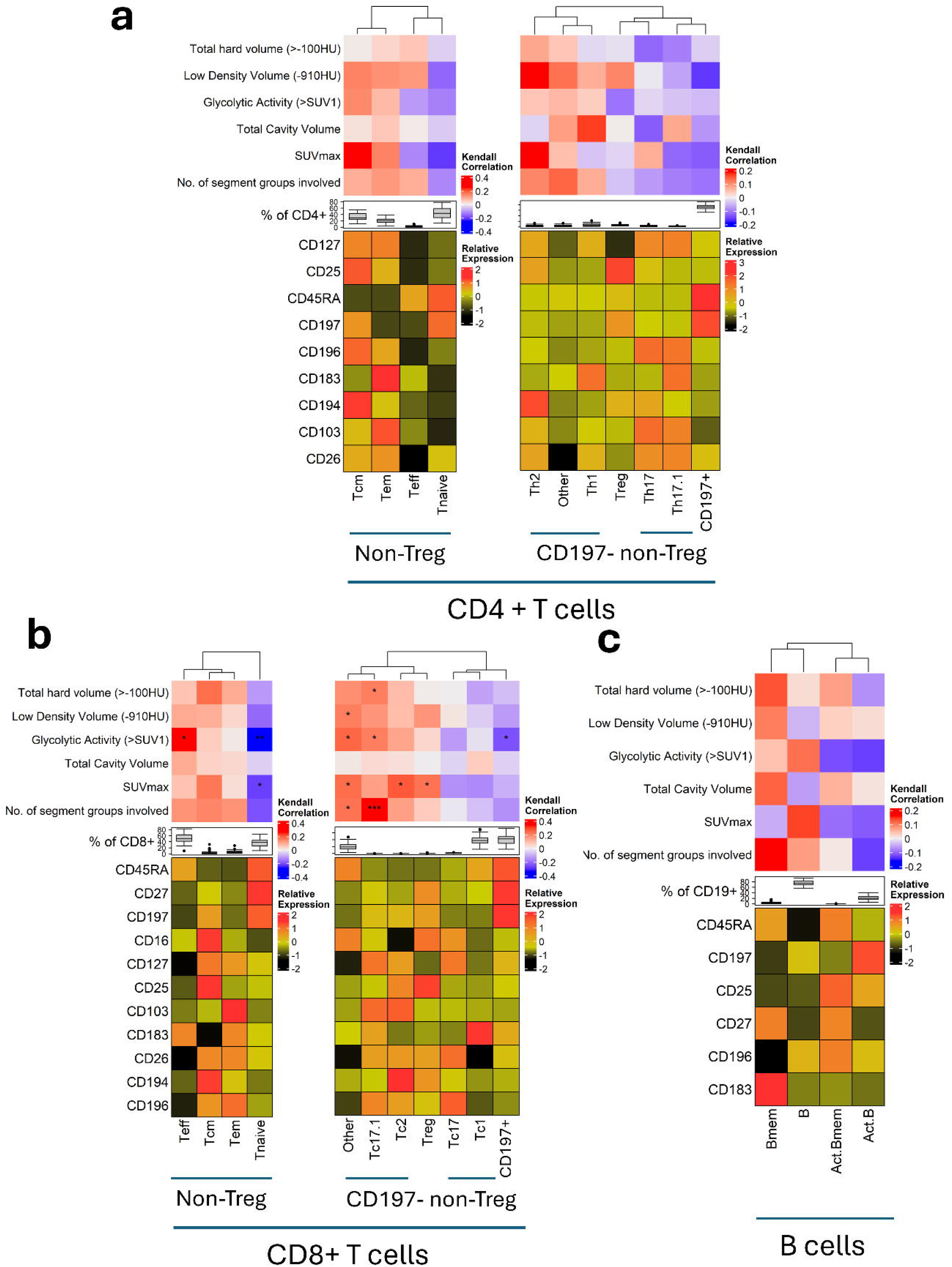
Heatmaps displaying correlations of cell phenotypes with clinical features of lung disease. Cells were clustered according to their expression of the markers shown in each expression heatmap. Metaclusters were annotated according to expected phenotypes for memory and functional subsets. Frequencies of each subset were correlated to clinical features of pulmonary function and disease using Kendall’s rank correlation test. The frequency of each subset among the total cell population across all participants is shown as boxplots. The median expression of markers used in clustering was scaled and is shown for each metacluster, defining cell function and phenotype; the range and median of expression was used to denote expression intensity in each expression heatmap. Figure 4a: CD4+ T cells. Treg were defined as CD25+CD127lo/- among total CD4+ cells. Memory phenotypes were inferred by the expression of CD45RA and CD197 among non-Treg CD4+ T cells. Subpopulations of effector T cells were identified by the expression of CD194, CD183, or CD196 among CD4+CD197-non-Treg cells. Figure 4b: CD8+ T cell subsets were inferred similar to the CD4+ T cell criteria, using the CD8+CD4-population of T cells. Figure 4c: B cells were clustered according to their expression of CD45RA, CD27, CD25, CD197, CD196, and CD183 among CD19+CD3-lymphocytes. Activation and memory phenotype was defined by the expression of CD25 for activation and CD27 for memory. Homing was inferred by the expression of CD197 for movement to lymph nodes and CD183 for homing to sites of inflammation.

CD8 T cells displayed similar results (Figure 4b). The naive CD45RA+CD197+ population displayed a negative correlation with extent of disease, while chemokine receptor-expressing effector cells (Teff and Tem) and central memory (Tcm) populations displayed positive correlations. Specifically, we observed more subsets expressing CD194 (Tc2), CD196 (Tc17.1), and CD25 (Treg), and conversely low percentages of CD183 (Tc1) expressing subsets among participants with more extensive lesions, suggesting that Tc17.1, Tc2, and Tcreg subsets may be associated with lung pathology. CD103 expression was also present among those CD8 subsets correlated with lung pathology. Altogether suggesting that activated effector CD8 T cells are primed for migration to the site of inflammation.

Percentages of B cells (Figure 4c) expressing CD25 and CD196 (denoted activated B cells) were lower among participants with severe lung pathology. These B cells are also CD183-, suggesting they are less migratory to inflammatory sites. Conversely, memory B cells expressing CD45RA, CD27 and CD183 were positively correlated with some features of involvement.

Results of the sensitivity analysis are shown in the Supplementary materials.

## DISCUSSION

In this exploratory analysis of the immune correlates of functional, anatomical and metabolic abnormalities early after successful TB treatment, we have shown that abnormalities on pulmonary function tests and FDG PET-CT were common, but highly variable in both pattern and severity; that, while there was general consistency between measures of imaging and pulmonary function test severity in an individual, they were not correlated with measures of exercise tolerance and symptoms/quality of life, suggesting that PTLD is often subclinical; that markers of worsening severity of lesions on FDG PET-CT were reflected by higher peripheral blood concentrations of proinflammatory cytokines, and reduced regulation of inflammation by T helper cells; and that BALF markers of leukocyte trafficking, collagen degradation and remodelling, regulation and repair at the site of disease correlated with worsening severity of both FDG PET-CT and lung function tests.

We started our analysis in search of distinctive patient phenotypes, but rather than clustering according to spirometry classes or the morphology of anatomical abnormalities, we found that markers of these disease processes occurred together and were correlated to the number of lobes involved and to each other. The pathology could not be classified into separate phenotypes analogous to that found in other obstructive or fibrotic pulmonary diseases. As a result, this study is the first to suggest that it is was rather the norm for all these processes to be present in each individual, and greatly influenced by the radiologic extent of lung involvement. We suggest that in PTLD, lung function is reduced through simultaneous processes: 1) destruction of total lung capacity due to cavitation, bullae formation, bronchiectasis, and mass lesions with a large fibrotic component (which may further reduce capacity through restriction), 2) airflow obstruction with increased residual volume attributable to dysfunction of both large and small (distal) airways leading to further reduction of FVC, and possibly 3) more isolated vascular involvement that impairs diffusion.

We are also the first to show immunological evidence of dysregulated inflammation, ongoing on both systemic and organ-specific levels after TB cure. We measured proteins in serum and BAL fluid that implicated Type 1, Type 2, and regulatory responses, as well as ongoing remodelling of lung tissue, airways, and vasculature. Through flow-cytometry, we also found an association between extent of lung pathology and subsets of CD8 and CD4 T-cells in keeping with a response to ongoing pro-inflammatory signals and antigenic stimulation in tissue and the periphery, namely central memory, and effector populations primed for recruitment in the periphery. In contrast, we found a lack of activated memory B cells primed for movement to the site of disease. The dysregulation and chronic stimulation of inflammatory cells will likely be detrimental to repair processes and decrease effective immune responses against pathogens. We found some promising markers of impairment like SP-D in serum, and the number of lobes affected as a basic radiographic measure of extent which could potentially be derived from chest x-ray analysis as well.

Our study was exploratory and limited by a small sample size and cross-sectional design. Sensitivity analysis showed a loss of significance when data subsets were left out of analysis. As such, individual correlations should be interpreted with caution. Further limitation was introduced by the inherent variability of BAL fluid results, as protein levels reflect only the area sampled and there is no perfect normalisation method.

## Conclusion

In TB survivors, there is correlation between the extent of structural damage and pulmonary function impairment. In turn, this influences both local and systemic inflammatory responses towards dysregulation and ongoing remodelling. Further work is required to find ways to prevent, identify, measure, and treat PTLD.

## Acknowledgements

We thank CRDF for funding support through grant number: This publication is based on work supported by a grant (G-202212-69613) from the U.S. Civilian Research & Development Foundation (CRDF Global). Any opinions, findings and conclusions or recommendations expressed in this material are those of the author(s) and do not necessarily reflect the views of CRDF Global.

Sincere gratitude to the participants for their willingness to participate and comply to study procedures. We further acknowledge the Regional Prospective Observational Research in Tuberculosis (RePORT) International and RePORT South Africa consortia for their partnership. We thank Francois Swart and his skilled team of lung technologists at Tygerberg Hospital, as well as Dr Alex Doruyter and his team at Numeri Node for Infectious Imaging.

## Data Availability Statement

The data that support the findings of this study are available from the corresponding author upon reasonable request.

## Human/Animal Ethics Approval Declaration

This study was performed in accordance with the Declaration of Helsinki. This human study was approved by the Stellenbosch University Health Research Ethics Committee - approval: N22/11/135 and by the University of Cape Town Human Research Ethics Committee - approval: 004/2023. All adult participants provided written informed consent to participate in this study.

