## Supplementary materials for "Immune, metabolic, anatomical, and functional features of people after successful tuberculosis treatment: an exploratory analysis"

### 1. Supplementary methods

**Recruitment and sample collection:**

We obtained Ethics approval for the study from SU HREC, reference number N22/11/135 and UCT HREC reference number 004/2023. Adults who were within 6 months after successful treatment for drug sensitive TB were screened and enrolled after informed consent. We excluded those who recently used systemic corticosteroids, had current active infection other than HIV, with known or suspected malignancy, were pregnant or breastfeeding, or had untreated HIV. We performed recruitment in January 2023, until May 2023 in areas that included Cape Town and the Western Cape Winelands (peri-urban and rural towns of Paarl, Worcester, Ceres, Robertson, Rawsonville, Wellington, De Doorns). We took medical history, performed directed physical examination and performed side room investigations including rapid HIV-testing, haemoglobin, random glucose, and point-of-care HbA1c. We then collected blood, sputum, and nasopharyngeal swabs for SARS-CoV-2 PCR. Active TB was excluded by sputum auramine smear microscopy and liquid culture (BACTEC MGIT, Becton Dickinson, USA), as well as clinical assessment. We also tested for Aspergillus activity on sputum PCR. Participants' study visits and procedures were completed within a 4-week window. One asymptomatic participant's nasal swab tested positive for SARS-Cov-2 on Xpert Xpress which necessitated re-scheduling study procedures until after potential infectiousness. We also found that four participants' (all of whom had 4 or 5 lung lobes affected on imaging) post-BALF sputum tested PCR positive for Aspergillus.

**Clinical procedures:**

A questionnaire specifically for post-TB lung disease has not been validated. Therefore, the Chronic Obstructive Pulmonary Disease (COPD) Assessment Test (CAT) and the St George's Respiratory Questionnaire (SGRQ) were identified as the most appropriate questionnaires for this patient population to test the subjective impact of lung health on enrolled participants[1–3]. These questionnaires were completed by the participants in their preferred language. Questionnaires were made available in English, Afrikaans and isiXhosa. A trained study nurse explained how to complete these prior to completion and assisted with the completion only when required due to low literacy levels of some participants.

Pulmonary function testing (PFT) was performed in a dedicated lung function laboratory at Tygerberg Hospital, Cape Town, South Africa, by a qualified pulmonary clinical technologist on a CareFusion Masterscreen Pulmonary Function Test and Body system (Jaeger, Würzburg, Germany) with JLab Lab Manager software V5.32.0.5. Participants had routine spirometry with forced vital capacity (FVC), forced expiratory volume in the first second (FEV1) in litres, FEV1 to FVC ratio, and measures of flow in distal airways; diffusing capacity of the lungs for carbon monoxide ($D_LCO$) measured through a single breath-hold technique reported as ml/min/mmHg then corrected for alveolar volume (VA) measured by helium diffusion, haemoglobin, and carboxyhaemoglobin levels measured in arterial blood; and body plethysmography reported in litres. All procedures were performed in accordance with current European Respiratory Society (ERS) and American Thoracic Society (ATS) standards [4–6]. Results were reported numerically and expressed as Z scores according to the Global Lung Initiative reference equations, using the recommended category for the South African population[7,8] Spirometry, $D_LCO$, and lung volume results were further interpreted according to current ERS/ATS recommendations[9] . The six-minute walking test (6MWT) was conducted on a 30 m flat indoor surface with intermittent peripheral oxygen saturation measurements, in accordance with the current ERS/ATS guidelines[10]. Results of the 6MWT were reported as distance walked in metres, and the lowest recorded saturation. A five-times-sit-to-stand test (5TSTS) was performed by the research worker who assessed the participant, by recording the time in seconds that the participant needed to stand up and sit down five times from a seated position on a straight-backed chair[11].

PET-CT imaging was performed at the NuMeRI Node for Infection Imaging (Central Analytical Facilities, Stellenbosch University), using a Vereos Digital PET with 64- channel (128-slice equivalent) CT scanner (Philips Medical Systems). Participants were fasted for 4–6 hours prior to the administration of fluorine-18 fluorodeoxyglucose (FDG). The radiopharmaceutical was produced either by NRF iThemba LABS (Gluscan®) or by PET Labs Pharmaceuticals (FDG-scan) and administered at 2.8 MBq per kg (±10%), with minimum activity 185 MBq (5 mCi) and maximum of 257 MBq (7 mCi), 55-65 minutes before commencement of the PET. The CT component of the scan included anterior and lateral surviews for positioning, a low-dose CT for attenuation correction and a high-resolution CT acquisition (kVp of 100, mAs of 155, rotation time 0.5s, pitch 0.89, collimation of 64x0.625). The combined maximum calculated effective radiation dose (including PET radiopharmaceutical and CT) was <10.4 mSv (<1.04 REM) per imaging examination. Clinical reports were provided based on consensus between a nuclear medicine physician and a radiologist.

Analytic reads were performed on MIM Software (v. 6.6 or higher, MIM Software Inc, Cleveland, Ohio, USA) by a single reader, with quality control performed by a second reader. CT-based measurements included 1) cavity volumes by using a 'region-grow' tool from a user-defined point, 2) cavity wall thickness at the widest level of the cavity, 3) volumes of very high density lung lesions corresponding with TB lesions and fibrosis using a -100 Hounsfield Unit threshold[12], very low density volumes that include air-trapping and emphysema using a threshold of >-910 Hounsfield Units[13], as well as the values for the kurtosis and skewness of the density in the lung region that should correspond to level of fibrosis– for the measured volumes based on thresholding, volumes of interests were drawn (segmented) that includes both lungs, prior to creation of subsegments based on the various thresholds of interest. We decided on the -910HU threshold for low density lesion over the alternative -950HU threshold after visualising volumes of interest for both and finding the latter not sensitive enough. We also captured the presence of morphological lesion types (small nodules, large nodules, bullae, consolidation fibrotic scars, bronchiectasis) per segment group as qualitative assessment. The segmented lung volumes of interest were also used for measuring PET-based metrics. First a subsegment was created of the area of the lung with increased FDG-avidity using SUV (standard uptake value) 1, as a cut-off. We then captured the total glycolytic activity in that volume of interest. The most intense uptake in the lung volume of interest (SUVmax) was also measured, and determined to be higher than mild if more avid than a reference value in the aortic blood pool.

Bronchoscopy was performed for the collection of broncho-alveolar lavage fluid (BALF) only on participants who were deemed fit for bronchoscopy, based on clinical state and test results. Bronchoscopies were performed under conscious sedation in a dedicated bronchoscopy suite by trained staff according to our standardized operation procedure as previously described[14,15] and the British Thoracic Society's bronchoscopy guidelines[16].

Data were directly captured via electronic data entry using Research Electronic Data Capture (REDCap 14.1.0 - © 2024 Vanderbilt University)[17,18] with the server located at Stellenbosch University. Access is user-specific and password controlled.

**Laboratory procedures:**

<u>Additional background to marker selection</u>

Previous studies have shown that cytotoxic and type-2 cytokine production by CD4 and CD8 T cells predominate in cavitary TB patients, however the directionality of this association is still debated [19,20]. Others suggest that this is typically a Th1 and Th17-driven response [21–25] associated with high levels of tumour necrosis factor (TNF)α, interleukin (IL)-6, and IL-1β, where infected macrophages die through necrosis rather than apoptosis [26] and there is an influx of neutrophils.

The pathological manifestations of fibrosis take on diverse forms. These include constricting lesions, associated with loss of parenchymal volume with or without surrounding traction effects (e.g. cicatricial bullae), but also thickening or dilation of airways, causing bronchiectasis, associated with infiltrating neutrophils, macrophages and T-cells [27] and pleural thickening, associated with higher levels of pleural fluid TGFβ [28].

Sample processing and storage

*PBMC and serum*

Venous blood samples were collected, transported on ice and processing begun within 4 hours of collection. Serum tubes stood upright for a minimum of 30 minutes, at room temperature, after blood draw to ensure complete clot formation. They were then centrifuged at 1200g for 10 minutes before they were aliquoted for storage at -80℃. PBMCs were isolated from heparinized tubes with Ficoll density gradient medium and centrifugation, with two washing steps using phosphate buffered saline (PBS) and an optional red cell lysis step with sterile 1x ACK buffer working solution (ThermoFisher) if there was visible red cell contamination. Cells were counted using the trypan blue exclusion method, resuspended in cryomedia [fetal bovine serum (FBS) with 10% dimethyl sulfoxide] and stored in -80°C overnight in a Mr Frosty/CoolCell, then moved to liquid nitrogen the next day. For the assays, PBMCs were thawed in a water bath at 37°C until a small ice chip remained in the vial, then warmed media (RPMI with 20% FBS, 1 % L-Glutamine and sodium pyruvate) added to the vial in a dropwise fashion. The whole sample was then added slowly into a vial of warmed media, washed twice with media and centrifugation at 300g for 8 min, then rested overnight at 37°C in a concentration of $5x10^6$ cells per milliliter in a non-treated 96 well plate. Cells were then combined providing $3x10^6$ cells per participant for staining.

*BALF and cells*

BALF was placed on ice immediately after collection and processing begun within 2 hours. Aliquots were pooled while being filtered through a 100µm cell strainer, then centrifuged at 300g at 4°C for 10 min. BAL-fluid was removed and stored in aliquots of 4.5ml, slowly cooled and placed in the -80°C freezer for long term storage. The BALF-cell pellet was washed twice by cold centrifugation at 300g for 10 min with 10ml of media (RPMI 1640 with 1% L-glutamine containing 5% heat inactivated pooled human type AB serum, plus 100 IU/ml penicillin and 100 µg/ml streptomycin and Fungizone-Amphot), with an optional red cell lysis step using sterile 1x ACK buffer working solution (ThermoFisher) if there was visible red cell contamination. Cells were then resuspended in media and counted by Trypan blue exclusion. An additional 10 µL cells in RPMI with 1ml PBS were prepared on glass slides with Cytospin, fixed with RapiDiff fixative, sequentially stained with acidic and basic dye, then washed once with PBS for cell differential count. Remaining BALF cells were resuspended in cryomedia after counting, placed into a Mr Frosty/Cool Cell and stored overnight in the -80°C freezer for next-day transfer to liquid nitrogen.

### Panel design and assays

#### Luminex®

Measurement of analyte concentrations. To assess the concentrations of cytokines and hormones in blood and BALF specimens, Luminex and ELISA were performed. BALF specimens were filter-sterilized using a 0.22uM syringe driven filter (GVS). BALF specimens were then freeze dried (Labconco) and resuspended in PBS to concentrate the specimens to 5:1. The custom Luminex panels were designed to include known markers of interest in other lung diseases/disease processes (fibrosis, cavitation, emphysema, bronchiectasis etc), markers which have been shown to have importance in PTLD (mainly MMPs), and standard markers of Type I, II and III inflammation and regulation. Custom designed Luminex kits (LXSAHM; R&D Systems) were acquired and assayed on serum and BALF specimens according to manufacturer instructions.

Briefly, samples were thawed, vortexed and centrifuged, then diluted according to the specific kit recommendations. The kits' standards, microparticle beads, detection antibody and Strepavidin-PE were reconstituted according to the kit guidelines and the Luminex assay was performed. Assays were read on both the Bio-Plex 200 platform where the Bioplex Manager software 6.2 was used for bead acquisition and the xMAP Intelliflex instrument which used the xPonent 4.2 analysis software for bead acquisition. The Bio-Plex Manager software 6.0 was used for sample analysis.

Luminex analytes included CCL3/MIP-1α; CCL11/Eotaxin; CX3CL1/Fractalkine; CXCL9/MIG; CXCL13/BLC/BCA-1; EGF; FGF basic/FGF2/bFGF; G-CSF; Growth Hormone; IFNγ; IL-2; IL-4; IL-8/CXCL8; IL-10; IL-12/IL-23 p40; IL-13; IL-17/IL-17A; IL-21; IL-33; M-CSF; MMP-1; MMP-7; MMP-8; SP-D; TGFα; TNFα; VCAM-1/CD106; VEGF; E-Selectin/CD62E; CCL2/JE/MCP-1; IL-1β/IL-1F2; α2-Macroglobulin; CCL18/PARC; MMP-2; PDGF-AA; TIMP-1; Uteroglobin/SCGB1A1; Adiponectin/Acrp30. ELISA kits were used for CXCL12/SDF-1a (DSA00) and Cortisol (KGE008B) from R&D Systems; and for DHEA (NBP2-61256), TIMP3 (KA0420), and Albumin (NBP3-12186) from Novus Biologicals. The CXCL12/SDF-1a ELISA was performed on plasma specimens according to manufacturer's recommendations, while the remaining kits used serum specimens.

#### *Flow cytometry*

To assess the frequencies of immune cell subsets in the PBMCs of participants, flow cytometry was performed. Cryopreserved PBMCs were thawed and rested overnight at 37C in a non-treated flat-bottom 96 well plate at 5 million cells per ml. Cells of each participant were then combined and transferred to a 96 well round bottom plate. Up to 3 million cells were stained for each participant. Cells were stained with Live/Dead Violet fixable viability dye (Thermofisher), washed and fixed in Fixation Buffer (Biolegend).

Cells were then stained before and after permeabilization for anti-cytokine antibodies with pre-titrated volumes of IL-17A BV421 (clone N49-653; 562933), CD16 BV510 (clone 3G8; 563830), CD103 BV480 (Ber-ACT8; 746472), CD194 BV605 (1G1; 562906), CD26 BV650 (M-A261; 744451), CD196 BV711 (11A9; 563923), CD25 BV786 (M-A251; 563701), CD4 PerCP-Cy5.5 (SK3; 332772), CD197 BB790-P (624296), TNFα PE (6401.1111; 340512), CD14 APC-Cy7 (MφP9; 333951) and CD19 PE-Cy5 (HIB19; 555414) from BD Biosciences; CD3 Spark Blue 550 (SK7; 344852), CD127 (IL-1Ra) PE-Fire700 (A019D5; 351366), IFNy PE-Cy7 (4S.B3; 502528), CD45RA APC (HI100; 304112), CD27 APC-Fire810 (O323; 302864) from Biolegend; CD8 cFluor V547 (SK1; R7-20064) and CD56 cFluorR720 (5.1H11; R7-20089) from Cytek Biosciences; TGFβ Vio Bright FITC (CH6-17E5.1; 130-106-994) from Miltenyi Biotec; and CD183 (CXCR3) PE-eFlour610 (CEW33D; 61-1839-42) from Thermofisher. For surface and intracellular staining, cells were stained for 1 hour at 4C. Acquisition of stained cells was performed on a Cytek Aurora spectral flow cytometer (Cytek Biosciences).

*Flow Cytometry Data Analysis*. Acquired flow cytometry data was unmixed and compensated using SpectroFlo Software (Version 3.2; Cytek Biosciences). Unmixed data was gated to remove poor quality events, doublets, debris, and dead cells in FlowJo (v10.9.0; BD Biosciences). Channel values were exported from Flowjo separately for CD4+ T cells, CD8+ T cells, and B cells, and analysed further in R (R Core Team (2023). R: A Language and Environment for Statistical Computing. R Foundation for Statistical Computing, Vienna, Austria). All participant data and relevant markers were used to cluster within each data set using FlowSom (Van Gassen S et al. (2015) FlowSOM: Using self-organizing maps for
visualization and interpretation of cytometry data. Cytom Part J Int Soc
Anal Cytol 87: 636-645.) with 24 dimensions and the number of clusters set to 100. Clusters were then annotated based on expression levels of each relevant marker in accordance with the gating strategy. Expression of cytokines was based on relative expression within each cluster. Correlations between clinical parameters and cluster or cell subset frequencies were performed using Kendall correlation tests. Heatmaps were produced using the ComplexHeatmap package (Gu, Z. (2022) Complex Heatmap Visualization. iMeta).

**Statistical analysis:**

All statistical analyses were conducted using R version 4.3.3 [29]. Data clean-up was completed using R packages *tidyverse* v2.0.0 [30], *dplyr* v1.1.4[31], *data.table* v1.15.0 [32] and *tidyr* v1.3.1[31] and *reshape2* v1.4.4 [33]. Summary statistics consisting of the mean, median, minimum and maximum values were obtained using the *summary* function in base R. Data distribution and potential outliers was visualised using box and whisker plots drawn using the *ggplot2* package v3.4.4 [34].

Data was tested for normality using Shapiro-Wilks test with subsequent normalization of non-normal variables either through log transformation of the variable, taking the square root of the variable or calculating the cube of the variable. The majority of variables remained non-normally distributed despite transformation, and were analysed using non-parametric methods during correlation analyses.

Correlations within the data were explored using the R packages *corrplot* v0.92 [35], and *ppcor* v1.1[36] (for partial correlations), with visualisations through *ggpubr* v0.6.0[37] and *ggcorrplot* v0.1.4 [38]. For non-normally distributed data Kendall's Tau was used to assess correlation. Correlation within the data was assessed first by grouping the data into subsets of (a)PET-CT variables and CAT score, (b) St. George's Questionnaire scores and physical exercise tests, (c) lung function variables, (d) Luminex analytes from BAL and (e) Luminex analytes from SERUM and Flow cytometry results. Thereafter, the sub-data sets were compared all against all. For each correlation being made between two variables, samples that had missing data for the variable were omitted from the correlation.

Correlation to confounding socio-demographic and wellness features, such as age, BMI, years smoking and illicit drug use (list in supplementary table 3) were performed and if variables correlated significantly to any of these factors, partial correlation was performed, correcting for these features. To minimise the impact of multiple testing and forgo post-hoc correction, those correlations which were significant were assessed using sensitivity analysis. Briefly, ten subsets of the data, where 5% of the samples were randomly removed at every iteration, were tested for correlation between the significant variables. If a correlation remained significant, despite subsetting of the data, we concluded that no particular sample was driving the effect and that it was worth noting for future study. Instances where one or more data subsets showed a non-significant correlation of a previously significant interaction, we concluded that the missing samples from that subset were potentially driving the significant effect, or that the significant effect does not hold true under a smaller sample size and should be interpreted with caution.

### 2. Supplementary Tables

**Supplementary Table 1:** Luminex and ELISA kits to screen peripheral blood and bronchoalveolar lavage fluid protein concentrations.

| Assay | Kit Name | Cat. No. | Supplier | Specimen | Analytes |
|---|---|---|---|---|---|
| Luminex | Human Magnetic Luminex Screening Assay | LXSAHM-29 | R&D | Serum & BALF | CCL3/MIP-1α; CCL11/Eotaxin; CX3CL1/Fractalkine; CXCL9/MIG; CXCL13/BLC/BCA-1; EGF; FGF basic/FGF2/bFGF; G-CSF; Growth Hormone; IFNγ; IL-2; IL-4; IL-8/CXCL8; IL-10; IL-12/IL-23 p40; IL-13; IL-17/IL-17A; IL-21; IL-33; M-CSF; MMP-1; MMP-7; MMP-8; SP-D; TGFα; TNFα; VCAM-1/CD106; VEGF; E-Selectin/CD62E |
| Luminex | Human Magnetic Luminex Screening Assay | LXSAHM-02 | R&D | Serum & BALF | CCL2/JE/MCP-1; IL-1β/IL-1F2 |
| Luminex | Human Magnetic Luminex Screening Assay | LXSAHM-01 | R&D | Serum | α2-Macroglobulin |
| Luminex | Human Magnetic Luminex Screening Assay | LXSAHM-05 | R&D | Serum | CCL18/PARC; MMP-2; PDGF-AA; TIMP-1; Uteroglobin/SCGB1A1 |
| Luminex | Human Magnetic Luminex Screening Assay | LXSAHM-01 | R&D | Serum | Adiponectin/Acrp30 |
| Luminex | Human Magnetic Luminex Screening Assay | LXSAHM-01 | R&D | Serum | Fibronectin |
| Luminex | Human Magnetic Luminex Screening Assay | LXSAHM-07 | R&D | BALF | α2-Macroglobulin; Adiponectin/Acrp30; CCL18/PARC; Fibronectin; MMP-2; TIMP-1; Uteroglobin/SCGB1A1 |
| Luminex | Human Magnetic Luminex Screening Assay | LXSAHM-04 | R&D | Plasma | Galectin-1.Galectin-3, Galectin-9, Galectin-3 Binding Protein |
| ELISA | Human CXCL12/SDF-1 alpha Quantikine ELISA | DSA00 | R&D | Plasma | CXCL12/SDF-1α |
| ELISA | DHEA ELISA | NBP2-61256 | Novus Biologicals | Serum | DHEA |
| ELISA | Cortisol ELISA | KGE008B | R&D | Serum | Cortisol |
| ELISA | TIMP-3 ELISA | KA0420 | Novus Biologicals | Serum | TIMP-3 |
| ELISA | Human CXCL12/SDF-1 alpha Quantikine ELISA | DSA00 | R&D | BALF | CXCL12/SDF-1α |
| ELISA | DHEA ELISA | NBP2-61256 | Novus Biologicals | BALF | DHEA |
| ELISA | Cortisol ELISA | KGE008B | R&D | BALF | Cortisol |
| ELISA | TIMP-3 ELISA | KA0420 | Novus Biologicals | BALF | TIMP-3 |
| ELISA | Albumin ELISA | NBP3-12186 | Novus Biologicals | BALF | Albumin |

**Supplementary Table 2**: Fluorescent reagents used for flow cytometry panel on PBMC lymphocytes.

| Antibodies | | | | |
|---|---|---|---|---|
| Antigen | Fluorophore | Clone | Cat | Supplier |
| IL-17A | BV421 | N49-653 | 562933 | BD Biosciences |
| CD103 | BV480 | Ber-ACT8 | 746472 | BD Biosciences |
| CD16 | BV510 | 3G8 | 563830 | BD Biosciences |
| CD8 | cFluor V547 | SK1 | R7-20064 | Cytek Biosciences |
| CD194 | BV605 | 1G1 | 562906 | BD Biosciences |
| CD26 | BV650 | M-A261 | 744451 | BD Biosciences |
| CD196 | BV711 | 11A9 | 563923 | BD Biosciences |
| CD25 | BV786 | M-A251 | 563701 | BD Biosciences |
| TGFβ | Vio Bright FITC | CH6-17E5.1 | 130-106-994 | Miltenyi Biotec |
| CD3 | Spark Blue 550 | SK7 | 344852 | Biolegend |
| CD4 | PerCP-Cy5.5 | SK3 | 332772 | BD Biosciences |
| CD197 | BB790-P | 3D12 | Custom | BD Biosciences |
| TNFα | PE | 6401.1111 | 340512 | BD Biosciences |
| CD183 (CXCR3) | PE-eFlour610 | CEW33D | 61-1839-42 | Thermofisher |
| CD19 | PE-Cy5 | HIB19 | 555414 | BD Biosciences |
| CD127 (IL-1Ra) | PE-Fire700 | A019D5 | 351366 | Biolegend |
| IFNy | PE-Cy7 | 4S.B3 | 502528 | Biolegend |
| CD45RA | APC | HI100 | 304112 | Biolegend |
| CD56 | cFluorR720 | 5.1H11 | R7-20089 | Cytek Biosciences |
| CD14 | APC-Cy7 | MφP9 | 333951 | BD Biosciences |
| CD27 | APC-Fire810 | O323 | 302864 | Biolegend |
| Dyes | | | | |
| Live/Dead Violet Fixable Viability Dye | | L34963 | Thermofisher | |

**Supplementary Table 3:** Confounders corrected for prior to correlation analysis and the variables they are associated with.

| **Variable** | **Confounders** |
|---|---|
| Total lung volume (mL) | sex, elicit drug use |
| Low density vol (< -910HU) | bmi, number of previous TB episodes, elicit drug use |
| Volume of low density | bmi, number of previous TB episodes, elicit drug use |
| FEV1 | bmi, number of previous TB episodes, |
| FEV1FVC | bmi, number of previous TB episodes, years smoking, elicit drug use |
| FEF2575 | bmi, number of previous TB episodes, |
| FEF75 | bmi, number of previous TB episodes, elicit drug use |

| | |
|---|---|
| FRV | bmi |
| Residual volume best | bmi, elicit drug use |
| DLCO (VA corrected) | elicit drug use |
| 6 minute walk | elicit drug use |
| Adiponectin in serum | bmi |
| MMP2 in serum | ethnicity |
| PARC in serum | age |
| PDGF in serum | bmi |
| TIMP in serum | ethnicity, bmi |
| Uteroglobin in serum | ethnicity, number of previous TB episodes |
| MMP1 in serum | bmi, sex |
| IL10 in serum | ethnicity, number of previous TB episodes |
| EGF in serum | ethnicity, elicit drug use |
| IL2 in serum | ethnicity |
| E-selectin in serum | ethnicity, years smoking, sex |
| SP.D in serum | years smoking, age, sex |
| TGFα in serum | ethnicity, elicit drug use |
| CCL11 in serum | age |
| CCL2 in BAL | age |
| Adiponectin in BAL | age |
| CCL18 in BAL | age, ethnicity |
| Fibronectin in BAL | age, bmi, ethnicity, elicit drug use |
| MMP2 in BAL | age |
| TIMP1 in BAL | bmi, ethnicity |
| Uteroglobin in BAL | bmi, sex |
| IFNy in BAL | years smoking |
| TNFα in BAL | age |
| IL8 in BAL | number of previous TB episodes |
| MMP1 in BAL | alcohol use, number of previous TB episodes |
| IL10 in BAL | age |
| EGF in BAL | age |
| VEGF in BAL | years smoking |
| CXCL13 in BAL | age |
| MIP1a in BAL | age, years smoking |
| IL4 in BAL | age, years smoking |
| IL17 in BAL | age |
| GH in BAL | age, years smoking |
| CX3CL1 in BAL | age, years smoking |
| CXCL9 in BAL | age, years smoking |
| VCAM in BAL | age |
| TGFα in BAL | bmi |
| IL13 in BAL | age |
| IL12 in BAL | age |
| CCL11 in BAL | age, years smoking |
| Albumin in BAL | bmi, ethnicity, elicit drug use |

| | |
|---|---|
| TIMP3 in serum | ethnicity |
| Cytotoxic T Cell | ethnicity, years smoking |
| Th1 | previous TB episode |
| Th17 | ethnicity, years smoking, alcohol use |
| Th2 | years smoking |
| Treg | ethnicity |
| Central memory T-Helper | years smoking, age |
| Naïve T-Helper | age |
| Large nodules | ethnicity, alcohol use |
| Small nodules | number of previous TB episodes |
| Cavity number | bmi |
| Fibrotic scarring | number of previous TB episodes |
| bullae | years smoking, age |

**Supplementary Table 4a: Cross-tabulation of spirometry measurements**

| | Spirometry Classifications | | | |
|---|---|---|---|---|
| | Normal FEV1 and FVC | Mixed (Low FVC and FEV1) | possible restrictive (Low FVC) | Obstructive Pattern (Low FEV1) |
| low FEF2575 | 15.38% | 100.00% | 77.78% | 83.33% |
| normal FEF2575 | 84.62% | 0.00% | 22.22% | 16.67% |

**Supplementary Table 4b: Cross-tabulation of diffusing capacity of the lungs for carbon monoxide and plethysmography classes.**

| | | Lung Volume Groups | | | | |
|---|---|---|---|---|---|---|
| | | Normal | Complex Restriction | Hyperinflation | mixed obstruction with restriction | Restriction |
| $D_LCO$ | normal | 70.00% | 0.00% | 33.33% | 50.00% | 100.00% |
| | loss of alveolar capillary structure with loss of lung volume | 0.00% | 60.00% | 25.00% | 100.00% | 0.00% |
| | localised loss of lung volume | 0.00% | 20.00% | 0.00% | 0.00% | 0.00% |
| | pulmonary vascular abnormality | 30.00% | 20.00% | 41.67% | 0.00% | 0.00% |

Complex restriction, TLC low; RV/TLC high; Hyperinflation, RV/TLC high; Mixed obstruction and restriction, TLC low; FEV1/FVC low; RV/TLC high; Restriction, TLC low; loss of alveolar capillary structure with loss of lung volume, Low $D_LCO$, Low VA; localised loss of lung volume, VA low; kCO high; pulmonary vascular abnormality, low $D_LCO$ only.

### 3. Supplementary Figures

**Supplementary Figure 1.** Flow of participants in the study

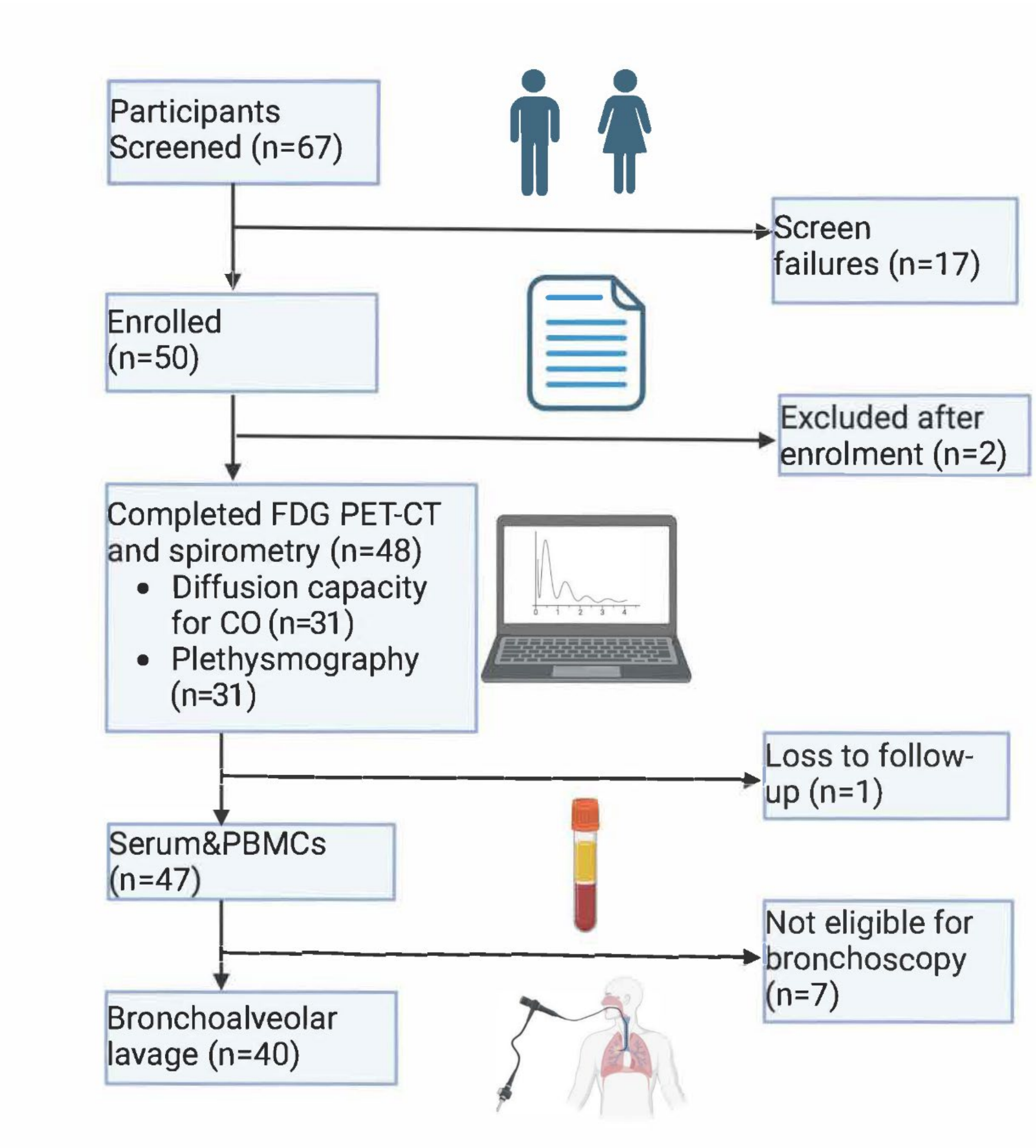

### 4. Results of the sensitivity analysis

Correlation matrix of quantified imaging variables, pulmonary function tests and participant characteristics to protein factors in serum (figure 3a). The following significant correlations failed to maintain significance during sensitivity analysis across one or more subsets of the data: High density vol(> -100HU) with IL-2, IL-8, E-selectin, SP-D, VEGF, PDGF, and MMP1; Cavity volume with SP-D, and uteroglobin; Glycolytic Activity (>SUV1) with TNFa, IL-12, CXCL9, CXCL12, E-selectin, SP-D, and VEGF; SUVmax with TNFa, IL-12, IL-4, E-selectin, SP-D, IL-10, VEGF, PDGF, TIMP-1, TIMP-3, and MMP1; FVC with SP-D, and uteroglobin; FEV1 with SP-D, uteroglobin, MMP1, and MMP8; FEF25/75 with, SP-D, PDGF, TIMP-1, and MMP1; Alveolar volume with PDGF, TIMP-1, MMP1, and MMP8; Years smoking with E-selectin, and SP-D; Age with SP-D; St Georges respiratory score with CCL2, Uteroglobin, VEGF, and TIMP-1.

Correlation matrix of quantified imaging variables, pulmonary function tests and participant characteristics to protein factors in bronchoalveolar lavage (Fig 3b). The following significant correlations failed to maintain significance during sensitivity analysis across one or more subsets of the data: Affected lobes with CCL18, and MMP2; High density vol(> -100HU) with Adiponectin; Cavity volume with CCL2, TIMP-1 and Adiponectin; Glycolytic Activity (>SUV1) with A2M, and Fibronectin; SUVmax with CCL18, E-selectin, and Adiponectin; FVC with IL-8, CCL2, and TIMP-1; FEV1 with IL-2, IL-8, Fibronectin, TIMP-1, and MMP2; FEF25/75 with IL-2, IL-33, IL-8, CCL18, A2M, and TIMP-1; Alveolar volume with IL-8, Uteroglobin, and MMP1; Years smoking with IFNy, IL-4, and CXCL13; Drug use with TNFa, IL-12, IL-4, IL-13, CCL2, CCL18, CXCL13, IL-10, Uteroglobin, VEGF, TIMP-1, and GH; Age with TNFa, IL-12, IL-4, IL-13, CCL18, CXCL13, IL-10, Adiponectin, and GH; St Georges respiratory score and CCL18.

Correlations of cell phenotypes with clinical features of lung disease (Figure 4). Cells were clustered according to their expression of the markers shown in each expression heatmap. Metaclusters were annotated according to expected phenotypes for memory and functional subsets. Frequencies of each subset were correlated to clinical features of pulmonary function and disease using Kendall's rank correlation test. The frequency of each subset among the total cell population across all participants is shown as boxplots. The median expression of markers used in clustering was scaled and is shown for each metacluster, defining cell function and phenotype; the range and median of expression was used to denote expression intensity in each expression heatmap.

10. Holland AE, Spruit MA, Troosters T, Puhan MA, Pepin V, Saey D, McCormack MC, Carlin BW, Sciurba FC, Pitta F, Wanger J, MacIntyre N, Kaminsky DA, Culver BH, Revill SM, Hernandes NA, Andrianopoulos V, Camillo CA, Mitchell KE, Lee AL, Hill CJ, Singh SJ. An official European respiratory society/American thoracic society

technical standard: Field walking tests in chronic respiratory disease. Eur Respir J. 2014;44(6):1428–46.

16. Du Rand IA, Blaikley J, Booton R, Chaudhuri N, Gupta V, Khalid S, Mandal S, Martin J, Mills J, Navani N, Rahman NM, Wrightson JM, Munavvar M. British Thoracic Society guideline for diagnostic flexible bronchoscopy in adults. Thorax. 2013;68(SUPPL. 1).

17. Harris PA, Taylor R, Thielke R, Payne J, Gonzalez N, Conde JG. Research electronic data capture (REDCap)-A metadata-driven methodology and workflow process for providing translational research informatics support. J Biomed Inform. 2009;42(2):377–81.

18. Harris M, Qi A, Jeagal L, Torabi N, Menzies D, Korobitsyn A, Pai M, Nathavitharana RR, Khan FA. A systematic review of the diagnostic accuracy of artificial intelligence-based computer programs to analyze chest x-rays for pulmonary tuberculosis. PLoS One. 2019;14(9):1–19.

19. Van Crevel R, Karyadi E, Preyers F, Leenders M, Kullberg BJ, Nelwan RHH, Van Der Meer JWM. Increased production of interleukin 4 by CD4+ and CD8+ T cells

from patients with tuberculosis is related to the presence of pulmonary cavities. J Infect Dis. 2000;181(3):1194–7.

20. Li S, Wang D, Wei P, Liu R, Guo J, Yang B, Zhang H, Lu J, Gao M, Pang Y. Elevated Natural Killer Cell-Mediated Cytotoxicity Is Associated with Cavity Formation in Pulmonary Tuberculosis Patients. J Immunol Res. 2021;2021:Article ID 7925903.

26. Stek C, Allwood B, Walker NF, Wilkinson RJ, Lynen L, Meintjes G. The immune mechanisms of lung parenchymal damage in tuberculosis and the role of host-directed therapy. Front Microbiol. 2018;9(October):2603.

27. King PT. The pathophysiology of bronchiectasis. Int J Chron Obstruct Pulmon Dis. 2009;4:411–9.

28. Sakamoto K, Kim MJ, Rhoades ER, Allavena RE, Ehrt S, Wainwright HC, Russell DG, Rohde KH. Mycobacterial trehalose dimycolate reprograms macrophage global gene expression and activates matrix metalloproteinases. Infect Immun. 2013;81(3):764–76.

29. R Core Team. R: A Language and Environment for Statistical Computing. Vienna, Austria; 2024.

30. Wickham H, Averick M, Bryan J, Chang W, McGowan LD, François R, Grolemund G, Hayes A, Henry L, Hester J, Kuhn M, Pedersen TL, Miller E, Bache SM, Müller K, Ooms J, Robinson D, Seidel DP, Spinu V, Takahashi K, Vaughan D, Wilke C, Woo K, Yutani H. Welcome to the {tidyverse}. J Open Source Softw. 2019;4(43):1686.

31. Wickham H, François R, Henry L, Müller K, Vaughan D. dplyr: A Grammar of Data Manipulation. 2023.

32. Barrett T, Dowle M, Srinivasan A, Gorecki J, Chirico M, Hocking T. data.table: Extension of `data.frame`. 2024.

33. Wickham H. Reshaping Data with the {reshape} Package. J Stat Softw. 2007;21(12):1–20.

34. Wickham H. ggplot2: Elegant Graphics for Data Analysis. Springer-Verlag New York; 2016.

35. Wei T, Simko V. R package “corrplot”: Visualization of a Correlation Matrix. 2021.

36. Kim S. ppcor: Partial and Semi-Partial (Part) Correlation. 2015.

37. Kassambara A. ggpubr: “ggplot2” Based Publication Ready Plots. 2023.

38. Kassambara A. ggcorrplot: Visualization of a Correlation Matrix using “ggplot2.” 2023.
